## Supplementary material for "Cutaneous surgical wounds have distinct microbiomes from intact skin": Tables 1-2, Supplementary Methods, and Supplemental Figures 1-7

##### Study Patients

The study was approved by the Partners Human Research Committee Institutional Review Board, and all subjects provided written informed consent prior to undergoing any study procedures. The skin microbiomes of patients at Massachusetts General Hospital undergoing MMS and managed by either complete or partial SIH were profiled 6-8 days after surgery. MMS is a clean procedure and pre-, intra- and postoperative antibiotics are not routinely administered. Postoperative wound care by patients was restricted to daily washing with soap and water, applying sterile petrolatum and a sterile non-adhesive bandage. Many patients were sampled at multiple different surgical sites. For each surgical site, anatomically-matched normal, intact skin site was sampled at the same time. The microbiome from each swab sample was then profiled using 16S rRNA sequencing of the V1-V3 region (1) and a computational approach enabling the classification of most skin bacteria at the species level (see Methods below). We collected samples from 70 patients; 5 of these were not analyzed because their sample was taken fewer than 6 or more than 8 days after surgery. Patients were excluded from downstream analyses if they received prophylactic antibiotics (7 patients) or had clinical evidence of surgical site infection at the one-week visit (2 patients). Samples were also filtered for quality control (see below); both matching control and surgical samples needed to pass quality control to be included in our analysis. If multiple pairs of wound and control samples from the same patient passed quality control, the site first sampled was used (to avoid patient-specific confounders). Three patients had no pair that passed quality control. A total of 53 pairs of surgical samples and controls were therefore included in our analysis (Table 1).

These samples were collected in two batches, the first of which contained swabs from the open surgical site and from the anatomically-matched site – either from the contralateral side or adjacent normal skin if surgery site fell along the sagittal plane, such as the dorsum nose – during routine clinical follow-up. These samples were processed in two sequencing runs, and are therefore referred to as batch 1A and 1B. To capture the microbiome on day of surgery as well as additional controls, a second study batch included additional swabs from the open wound and of the anatomically-matched site on day of surgery as well as at postoperative follow-up. Additional control swabs of the nares, alar crease, glabella, and shin were also obtained in batch two on the day of surgery. 18 swabs exposed to only air were also obtained as a negative control in both phases.

##### Sample Processing and Sequencing

All samples were obtained using sterile cotton swabs (PurFlock Ultra®) that were moistened with a drop of sterile saline before sampling. Sampled surfaces were rubbed using 40 brisk strokes, placed in a sterile container, and stored at -20°C until shipment to Microbiome Insights for processing and sequencing. Cohort characteristics are summarized in Table 1.

DNA extraction, sample prep, and sequencing were performed by Microbiome Insights (Vancouver, Canada). DNA extraction was performed using the MoBio PowerMag Soil DNA Isolation Kit. PCR was performed with dual-barcoded primers (2) targeting the 16S V1-3 (Bacteria) regions (27F: AGAGTTTGATCMTGGCTCAG, 534R: ATTACCGCGGCTGCTGG) for 35 cycles (Meisel 2016). The PCR reactions were cleaned up and normalized using the high-throughput SequalPrep 96-well Plate Kit and sequenced on the Illumina MiSeq to 300 cycles.

##### 16S Amplicon Analysis

Amplicon analysis was performed using the first 180 bp after the 27F primer using only the forward read. Cutadapt was used to trim and remove primers from reads (3), and QIIME2 version 2020.01(4) and DADA2 (5) were used to denoise raw reads using standard parameters, resulting in a table of amplicon sequence variants (ASVs) and their abundances across samples.

To classify 16S amplicon sequence variants (ASVs) at the species level, we first removed or relabeled errant sequences from version 132 of the SILVA database (6) using SATIVA (7), a program that checks the phylogenetic placement of entries against their taxonomic names. In particular, entries in the database from the genuses *Cutibacterium*, *Acidipropionibacterium*, *Pseudopropionibacterium* and families *Corynebacteriaceae* and *Neisseriaceae* were treated according to the following filters: (i) entries with incorrect higher taxonomic classes were removed; (ii) entries not classified at the species level were removed; (iii) entries that SATIVA (-x BAC) taxonomically classified with confidence below 90% were removed; (iv) entries that SATIVA identified as mislabeled with greater than 90% confidence were relabeled; and (iv) sets of species for which SATIVA identified >60% of entries as identical to another species were relabeled as a specific "taxa cluster". To reduce computational load, each family was run independently in SATIVA. Entries from the genus *Staphylococcus* were cleaned as in Khadka et al. (8). These processes removed about 2% of sequences from each group. The resultant quality-controlled database was used to train a naive Bayes classifier in QIIME2 (4).

Suspected contaminant ASVs from either environmental (including sample prep) or human DNA were identified and removed. For each batch, we identified ASVs that were significantly enriched in control samples taken from healthy skin relative to those from the other two batches combined and were therefore likely to be environmental contaminants (Wilcoxon Rank Sum, $p \leq 4 \times 10^{-5}$) (Supplementary Figure 1). This process removed 16 ASVs in total which made up an average abundance of 3.5% across samples; most of this abundance came from a taxa labeled as "Salinimicrobium: uncultured Pseudomonas" (3.1% in batch 3), which had a high match to chloroplast DNA when compared against NCBI's nucleotide database (blastnt -remote -db nt  -max_target_seqs 10 -word_size 30 -evalue 1e-60) (9, 10).

We also removed all ASVs labeled only "Bacteria", "Bacteria;Proteobacteria", or "Unassigned", as we suspected them to arise from human DNA. To confirm human origin of these ASVs, we compared a random subset of 100 ASVs from each of these groups against NCBI nucleotide database (blastnt -remote -db nt  -max_target_seqs 10 -word_size 30 -evalue 1e-60) (9, 10); the majority had their highest hit as human (94.9, 94.1, and 100%, respectively). Contrastingly, all other ASVs that classified only to the phyla matched bacterial isolates in NCBI's nucleotide database and were therefore retained. Lastly, several ASVs from the Gamma-Proteobacteria clade SUP05 were removed; this was marine bacteria included as positive control in all batches.

Relative abundances were then calculated by renormalizing using the remaining ASVs and counts. Samples with fewer than 500 remaining read counts after contamination removal were removed in downstream analyses.

**Statistical Methods and Phylogenetics**

For all comparisons between surgical and control sites, samples were only included only if both a surgical and matched control sampling site contained greater than 500 read counts after the sequencing depth filtering mentioned above. If a patient had multiple sampled sites that passed these filters, only the first site sampled was included in the matched-sample analysis to remove any patient-based bias. All statistical tests used are mentioned in the main text and figures where they are presented, using standard MATLAB functions and the FATHOM toolbox for distance metrics (11).

For phylogenetic reconstruction of *Corynebacterium* in Figure 2, the full 16S rRNA sequence of all *Corynebacterium* species found in our dataset was pulled from the SILVA database (version 132) (6). A phylogenetic tree was constructed in MEGAx using the pre-aligned sequences from SILVA and a neighbor-joining model with Tamara-Nei substitutions (12). This tree is rooted to the distant outgroup *Haemophilus massiliensis* (SILVA accession number HG931334).

### SUPPLEMENTAL TABLES:

Table 1. Characteristics of study participants and surgical sites

| Patient and Surgical Site Variables | Value |
|---|---|
| Demographics (n= 53 unique patients) | |
| Age, y, median (IQR) | 70 (18) |
| Sex, n (%) | |
| Male | 31 (58) |
| Female | 22 (42) |
| Race, n (%) | |
| Caucasion | 50 (94) |
| Unknown | 3 (6) |
| Ethnicity, n (%) | |
| Non-Hispanic | 48 (91) |
| Unknown | 5 (9) |
| Hispanic | 0 |
| Surgical Site Variables | |
| Anatomic location, n (%) | |
| Eyelid | 8 (15) |
| Postauricular/scalp | 6 (11) |
| Ear | 1 (2) |
| Nasal area | 15 (28) |
| Lip | 4 (8) |
| Forehead/temple | 9 (17) |
| Cheek/chin | 7 (13) |
| Neck | 2 (4) |
| Shin | 1(2) |
| SIH type and area, n (%) | |
| Complete SIH | 8 (15) |
| Area, $mm^2$, median (IQR) | 300 (528) |
| Partial Closure | 45 (85) |
| Area, $mm^2$, median (IQR) | 32 (31.5) |

*IQR = interquartile range

Table 2. : P-values for Comparison of Fractional Abundance between Wound and Matched Control Sites

| | With *Cutibacterium* | | | | *Cutibacterium* Removed | | | |
|---|---|---|---|---|---|---|---|---|
| **Genus** | **Mean Control Abundance** | **Mean Wound Abundance** | **Fold change on wounds** | **P-value*** | **Mean Control Abundance** | **Mean Wound Abundance** | **Fold change on wounds** | **P-value*** |
| *Cutibacterium* | 0.317 | 0.063 | 0.2 | $<10^{-6}$ | - | - | - | - |
| *Corynebacterium* | 0.181 | 0.365 | 2.0 | 0.001 | 0.253 | 0.396 | 1.6 | 0.013 |
| *Staphylococcus* | 0.255 | 0.375 | 1.5 | 0.088 | 0.377 | 0.392 | 1.0 | 0.940 |
| **Species** | | | | | | | | |
| *Cutibacterium acnes* | 0.307 | 0.060 | 0.20 | $< 10^{-6}$ | - | - | - | - |
| *Staphylococcus aureus* | 0.035 | 0.226 | 6.37 | 0.00 | 0.045 | 0.234 | 5.2 | 0.002 |
| *Staphylococcus epidermidis* | 0.146 | 0.114 | 0.78 | 0.04 | 0.223 | 0.120 | 0.5 | 0.004 |
| *Staphylococcus capitis* | 0.064 | 0.009 | 0.14 | $< 10^{-5}$ | 0.090 | 0.010 | 0.1 | $<10^{-6}$ |
| *Corynebacterium kroppenstedtii* | 0.107 | 0.059 | 0.55 | 0.002 | 0.147 | 0.064 | 0.4 | $<10^{-3}$ |
| *Corynebacterium tuberculostearicum* | 0.044 | 0.154 | 3.47 | 0.002 | 0.061 | 0.169 | 2.8 | 0.024 |
| *Corynebacterium accolens/fastidiosum* | 0.010 | 0.045 | 4.71 | 0.021 | 0.011 | 0.048 | 4.2 | 0.032 |

*Wilcoxon rank-sum test, not corrected for multiple hypothesis testing

### SUPPLEMENTAL FIGURES:

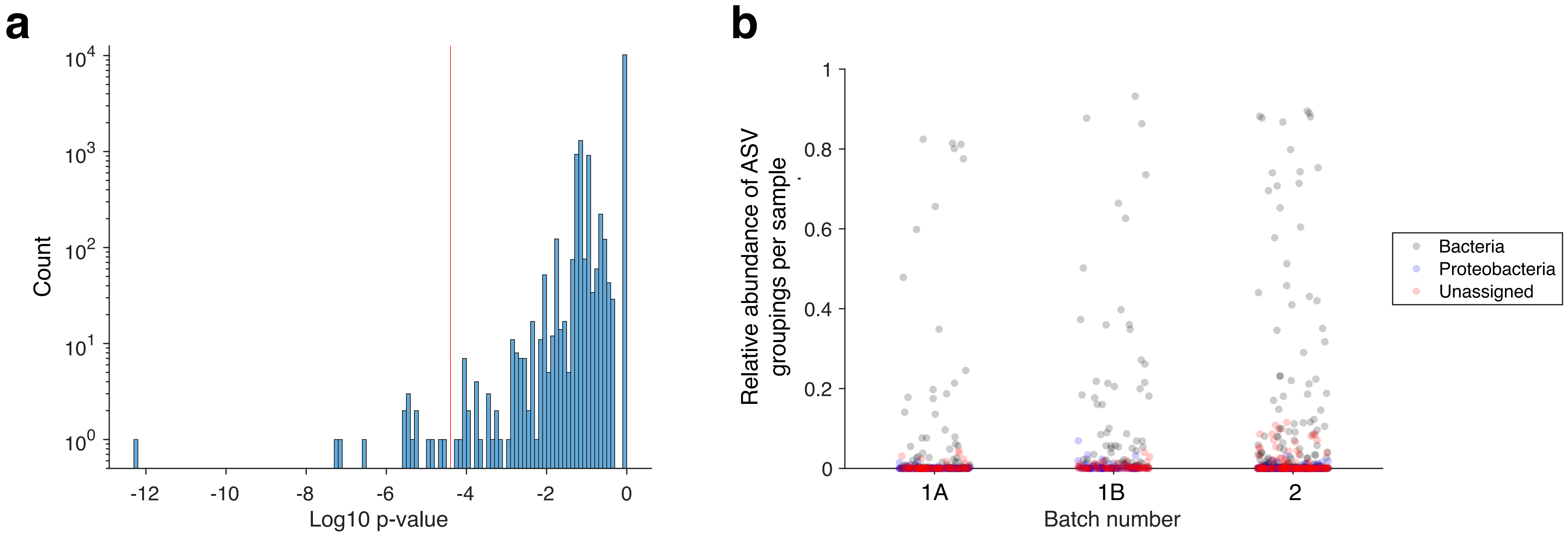


**Supplementary Figure 1: Evidence supporting removal of contaminant ASVs.** (a) Many ASVs showed significant differences between batches, suggesting contamination from the environment. For each batch and each ASV, we compared the relative abundance of ASVs between the focal batch and the other two batches combined via a Wilcoxon Rank Sum test. A histogram of the minimum p-value for each ASV (out of the 3 comparisons per ASV) is shown. The red line indicates the cutoff used for ASV removal (p ≤ 2.5 x 10-5). (b) ASVs classified by SILVA to the high taxonomic groupings of "Bacteria" and "Proteobacteria" or labeled as "Unassigned" were determined to largely arise from human DNA (Methods). For each of these labels and for each sample, we summed the relative abundance of all ASVs with that label. A scatter plot of these sums show that these ASVs made up a nontrivial amount of many samples. These ASVs were removed before subsequent analysis. This plot shows all samples, including those that did not have 500 reads after all filtering and were thus not included in downstream analysis.

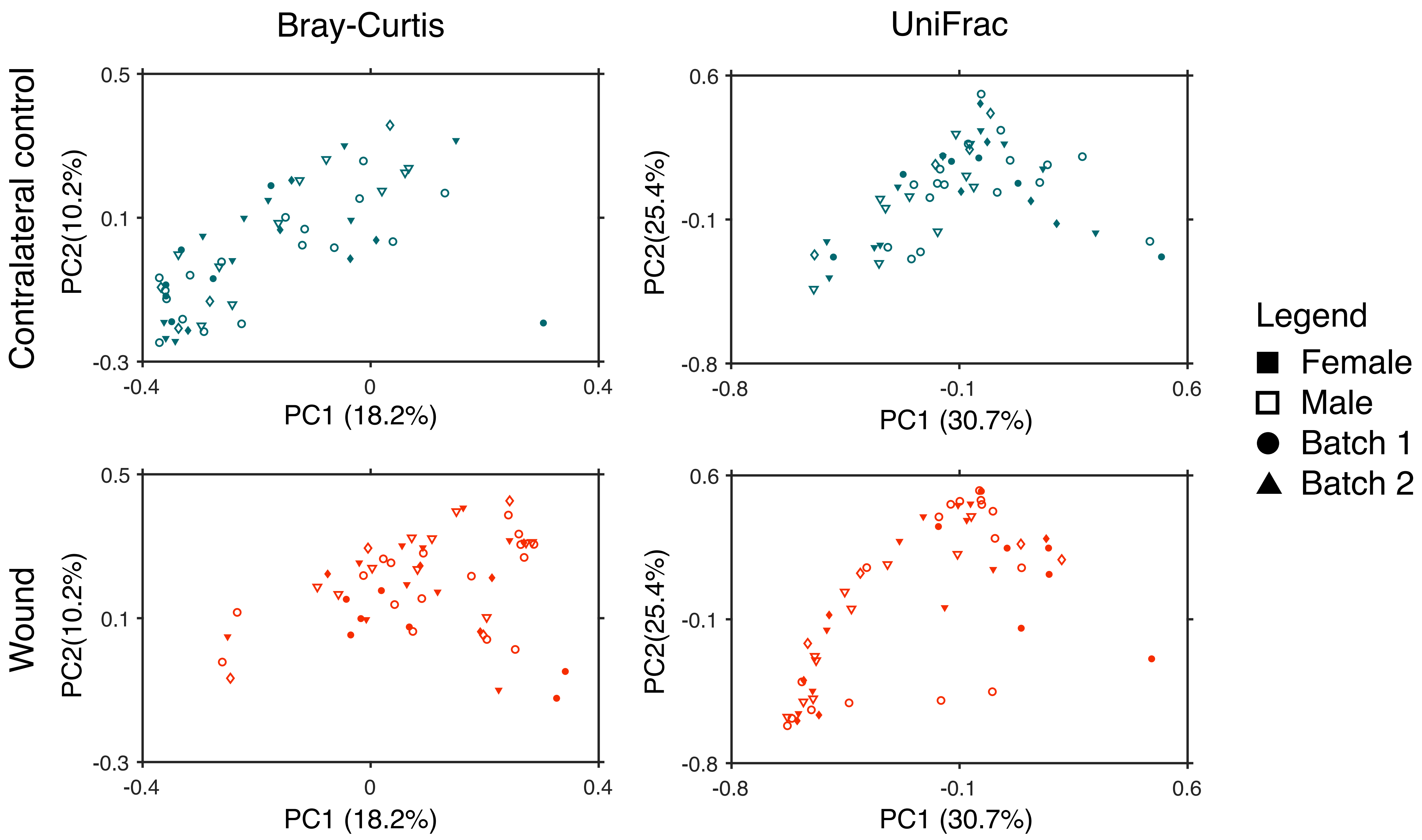


**Supplementary Figure 2: No bias is observed in microbiome distribution by sex.**

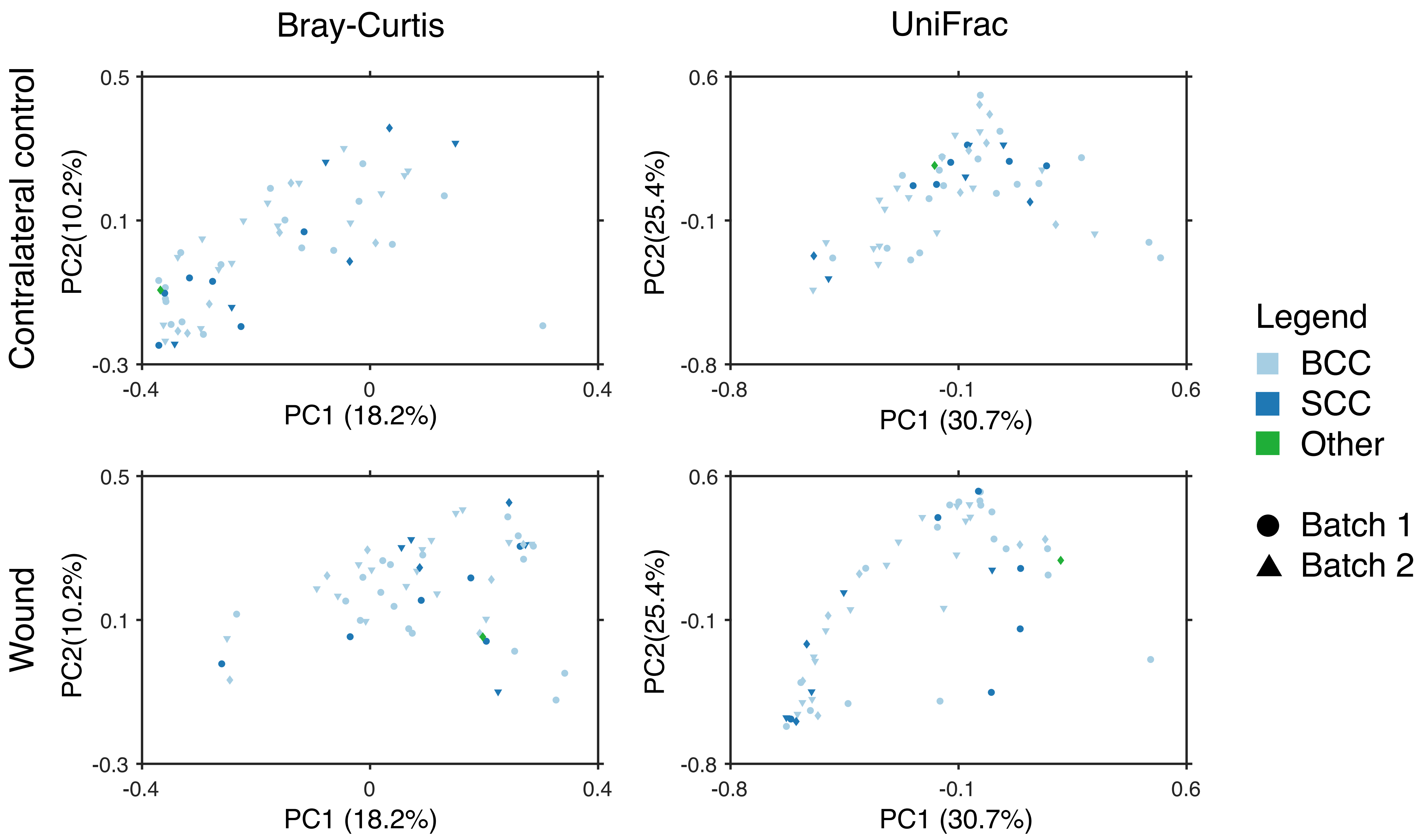


**Supplementary Figure 3: No bias is observed in microbiome distribution between cancer type.**

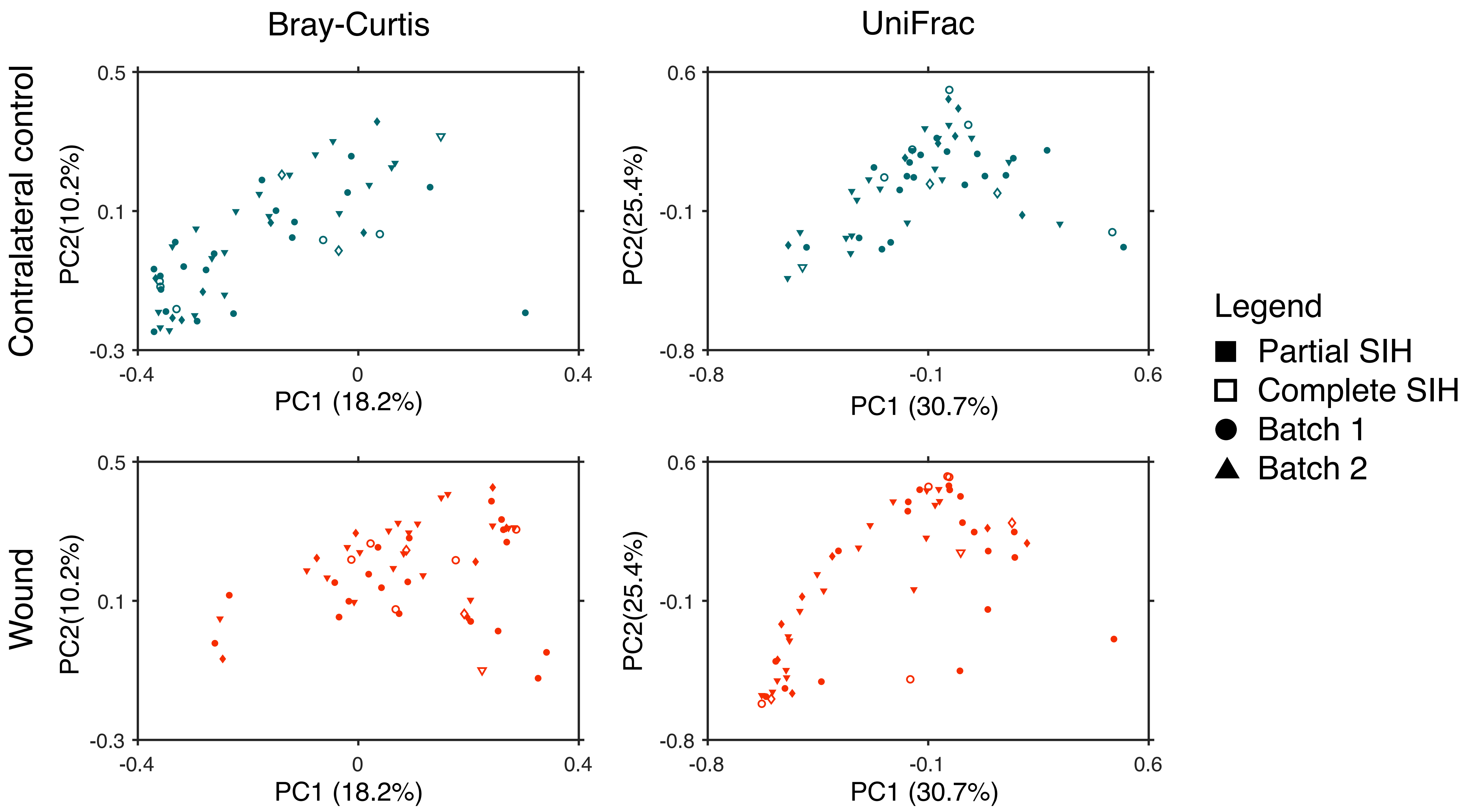


**Supplementary Figure 4: No bias is observed in microbiome distribution between wound closure type.**

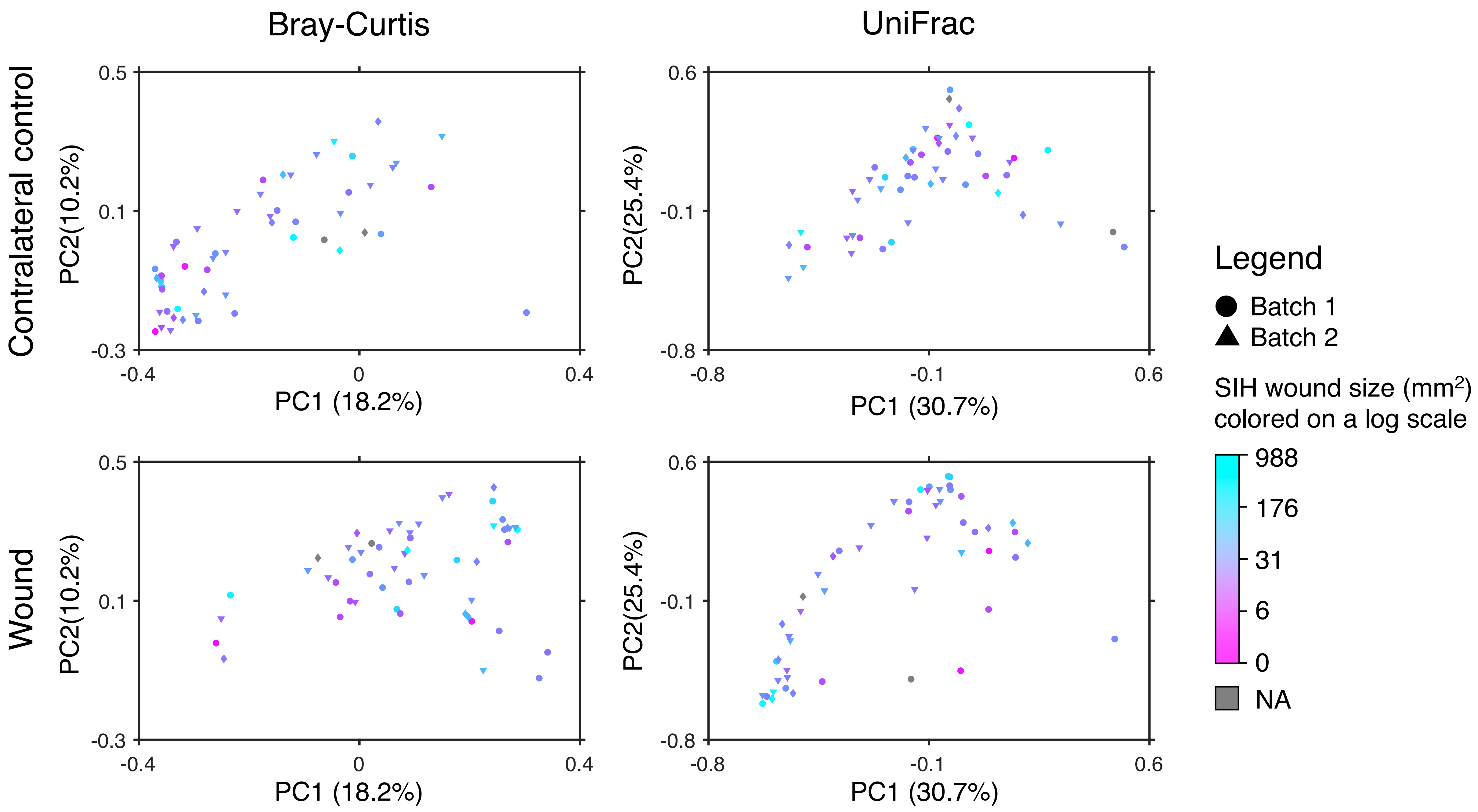


**Supplementary Figure 5: No bias is observed in microbiome distribution by wound size.**

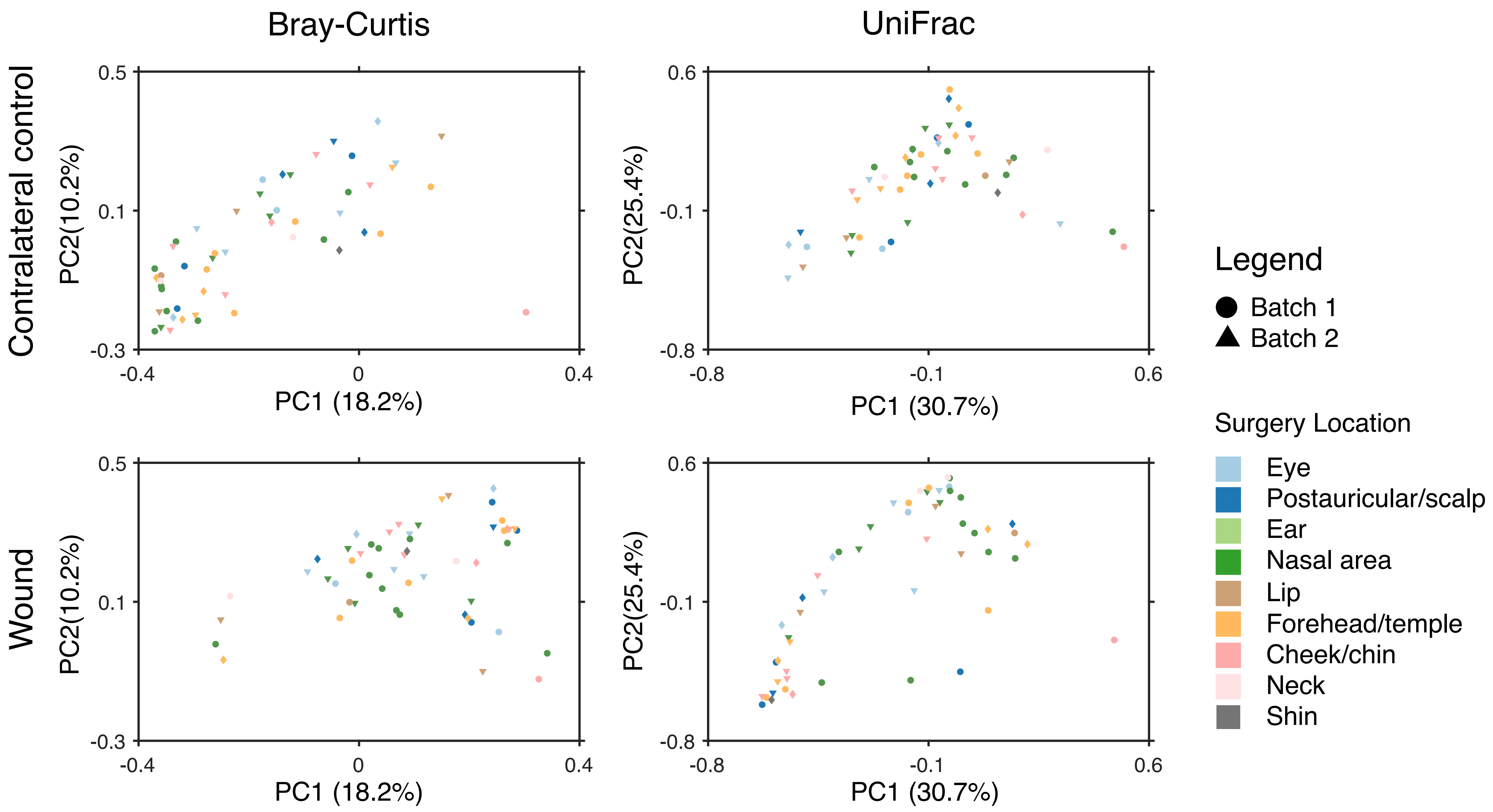


**Supplementary Figure 6: No obvious bias is observed in microbiome distribution by anatomical location.**

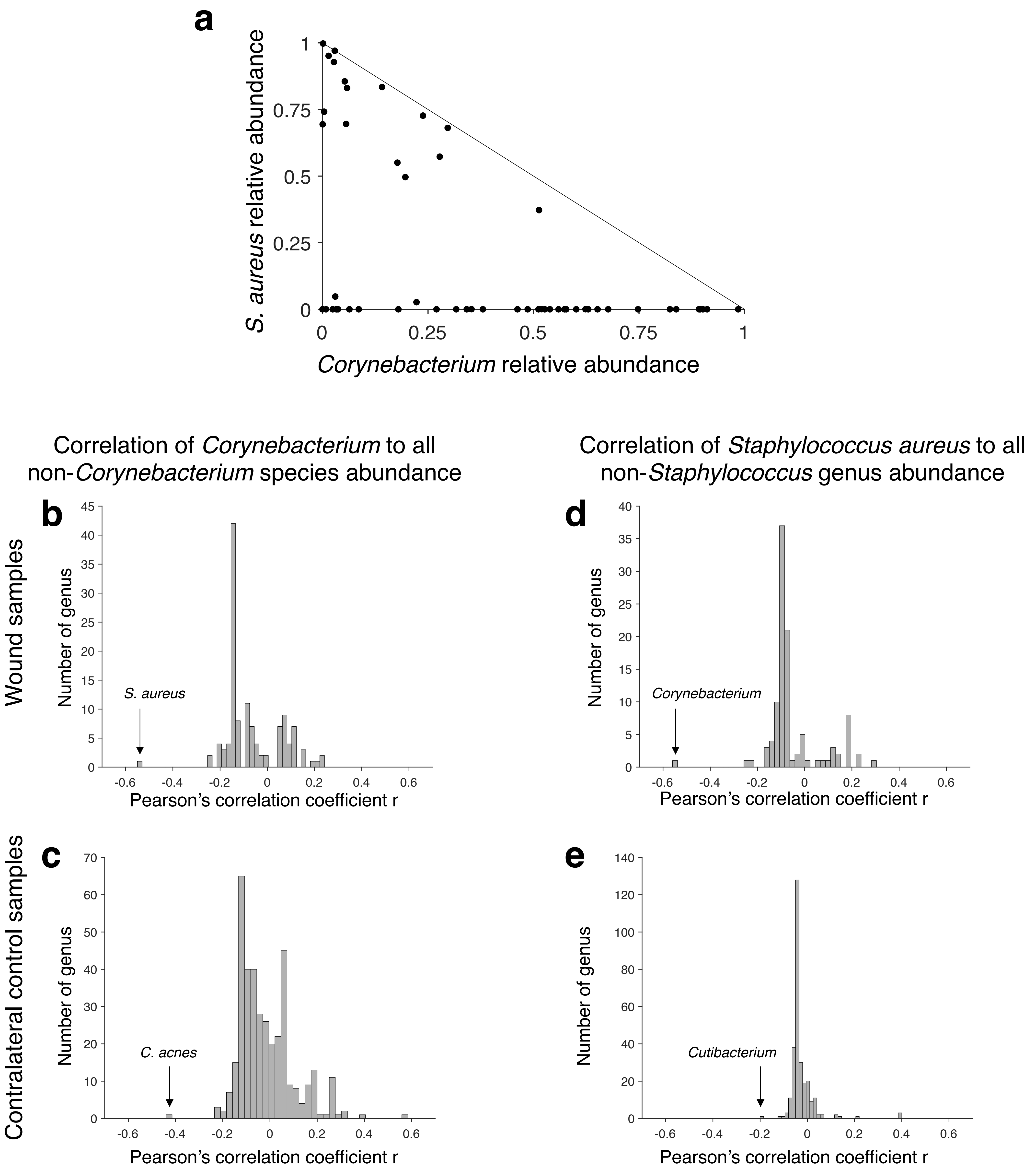


**Supplementary Figure 7: Comparing *Corynebacterium* and *Staphylococcus aureus* abundances reveals anti-correlation in wound samples.** (a) The relative abundances of *S. aureus* and *Corynebacterium* are negatively correlated in wound samples (R of -.55). (b) As these two taxa are the most abundant in skin, and therefore a negative correlation could emerge solely from the compositional nature of the data, we sought to contextualize this correlation. We calculated the correlation (Pearson's R) between the relative abundances of the *Corynebacterium* genus to all non-*Corynebacterium* species across all wound samples. *S. aureus* anti-correlates with *Corynebacterium* much better than any other species. (c) The same relationship is not true in control samples, where instead *C. acnes* correlates better with *Corynebacterium.* (d-e) A similar analysis in which the relative abundance of *S. aureus* is correlated against the relative abundance of all other genuses, again indicates that *Corynebacterium* uniquely correlates with *S. aureus* in a site-dependent manner. The difference in correlation strength between sites may not be a result in the difference of biological interactions at these sites; these taxa have very different relative abundance between sites, which can impact the strength of a detected correlation.
